# Clinical Presentation, Risk Factors, and Outcomes of Cardiovascular Disease in a Military Cardiac Center in Yemen

**DOI:** 10.1101/2025.08.04.25332803

**Authors:** Abdulelah Mansoor Al-Ganad, Ahmed Al-Motarreb, Taha Al-Maimoony, Nabil Al-Rabeei, Abdulaziz Aonallah, Esmail Al-Dabis

## Abstract

1.0

**Background:** Cardiovascular diseases (CVDs) are a growing public health concern in Yemen, yet detailed data from local facilities are limited. This study aimed to evaluate clinical characteristics, risk factors, and outcomes of CVDs in a military cardiac center.

**Aim:** To evaluate the clinical characteristics, risk factors, and outcomes of cardiovascular diseases (CVDs) among patients admitted to the Coronary Care Unit (CCU).

**Setting:** The study was conducted at the Military Cardiac Center in Sana’a, which includes a 7-bed CCU for cardiac cases, 75 general ward beds, 4 beds in the emergency department, and 2 beds in the operating theater.

**Methods:** A retrospective cohort study was conducted at the Military Cardiac Center in Sana’a, including all patients admitted to the Coronary Care Unit (CCU) in 2019. Clinical data were extracted from both electronic and archived paper records, including prior medical histories dating back to 2008 or 2010 when available. Variables included demographics, clinical presentations, diagnoses, prior cardiac history, and outcomes. Statistical analyses were performed using descriptive statistics, chi-square tests, ANOVA, and univariate logistic regression to identify predictors of in-hospital and 5-year mortality.

**Results:** A total of 446 patients were included (mean age:53.0 plus/minus 13.6 years; 84% male). The most common diagnoses were acute coronary syndrome (ACS, 60.9%), acute decompensated heart failure (ADHF, 13%), valvular heart disease (VHD, 9%), and cardiogenic shock (5%). Risk factors included khat use (76%), smoking (61%), hypertension (41%), diabetes (31%), and dyslipidemia (4.9%). Younger patients had higher rates of smoking, khat use, and prior myocardial infarction or PCI, while older patients had more diabetes, hypertension, and dyslipidemia. Significant predictors of in-hospital mortality included diabetes (OR=2.6), rheumatic heart disease (RHD, OR=2.4), cardiogenic shock (OR=11.3), heart failure (OR=9.2), sepsis (OR=31.1), and cardiac arrest (OR=74.0). For 5-year mortality, key predictors were unstable angina (OR=8.7), ACS (OR=4.0), elective PCI (OR=2.9), and khat use (OR=2.0).

**Conclusion:** CVDs in Yemen affect relatively young patients and are driven by modifiable, region-specific risk factors. Limited access to emergency interventions and late hospital presentation contribute to poor outcomes. Public awareness, preventive measures, and improved cardiac care infrastructure are urgently needed.

**Recommendations:** Enhancing public awareness, expanding access to cardiac emergency care, and strengthening preventive services are crucial to reducing mortality and improving outcomes. **Implementing these measures urgently can substantially mitigate the burden of cardiovascular disease in Yemen**.

## 2.0 Introduction

Cardiovascular diseases (CVDs)—including coronary artery disease, stroke, rheumatic heart disease, and peripheral arterial disease—remain the leading global cause of death. According to the World Health Organization, CVDs caused an estimated 17.9 million deaths in 2019, accounting for 32% of all global mortality **(1)**. Over 75% of these deaths occur in low- and middle-income countries (LMICs), where delayed diagnosis, limited access to specialized care, and prevalent behavioral risk factors—such as tobacco use, unhealthy diets, physical inactivity, and in Yemen, khat chewing—significantly contribute to the burden ***(1, 2)***.

Ischemic heart disease and stroke are responsible for nearly 87% of CVD-related deaths, many of which occur prematurely in individuals under age 70(3). In Yemen, CVDs account for over 33% of all deaths, making them the country’s leading cause of mortality (**4**) This crisis is exacerbated by ongoing conflict, economic hardship, weak infrastructure, and limited availability of evidence-based care. Modifiable risk factors— including khat use, malnutrition, and chronic stress—remain highly prevalent **(2)**.

While high-income countries have reduced CVD mortality through early detection and advanced care, Yemen still lacks robust data, particularly on critically ill patients in coronary care units (CCUs). Addressing this knowledge gap is essential for improving prevention and outcomes.

Therefore, this study aims to characterize the epidemiological profile, clinical features, risk factors, and outcomes of CVD patients admitted to the CCU at the Military Cardiac Center in Sana’a. By generating local evidence, the findings can inform targeted interventions and strengthen cardiac care within Yemen’s fragile healthcare system.

## 3.0 Objectives and Hypotheses of the Study

### 3.1 Study Objectives

#### 3.1.1 General Objective

To describe the clinical presentation, risk factors, course, and outcomes of cardiovascular disease among patients admitted to the Military Cardiac Center in Sana’a, Yemen.

#### 3.1.2 Specific Objectives

1. To determine the pattern of CVD among CCU-admitted patients.
2. To examine the distribution of CVD by demographic characteristics.
3. To identify major cardiovascular risk factors in the study population.
4. To assess hospital, stay duration, admission pathways, and treatment outcomes.
5. To compare the clinical presentation and outcomes of ACS between older and younger patients.
6. To identify predictors of in-hospital complications and mortality in high-risk cardiac patients.

## 4.0 Research Methodology

### 4.1 Study Setting

The study was conducted at the Military Cardiac Center in Sana’a, Yemen, a national referral hospital established in 2008. It includes a 7-bed Coronary Care Unit (CCU) staffed by cardiologists and nurses, and provides diagnostic and interventional cardiac services.

### 4.2 Study Design >

This retrospective cohort study included all patients admitted to the CCU in 2019. In addition to clinical data during admission, available past records— electronic or paper-based—were reviewed and entered when accessible. For most patients, prior medical histories were retrieved, in some cases dating back to 2008 or 2010. This enriched the dataset with baseline cardiovascular risk factors and comorbidities, strengthening the risk and outcome analyses.

### 4.3 Study Population

All adult patients (≥18 years) admitted to the CCU at the Military Cardiac Center in Sana’a during 2019 were included using total population sampling.

### 4.4 Inclusion and Exclusion Criteria

- **Inclusion:** Adults (≥18 years) admitted to the CCU in 2019 with complete cardiac records.
- **Exclusion:** Patients <18 years, early deaths (<6 hours), non-cardiac admissions, and incomplete data

### 4.5 Sample Size and Sampling Method

A total enumeration approach was used, including all eligible records from January to December 2019 identified via electronic and paper archives.

### 4.6 Data Collection Tools and Techniques

Data were collected using a structured checklist adapted from previous studies ***(5, 6)***.Variables included demographics, risk factors (smoking, khat use, diabetes, hypertension, dyslipidemia), clinical characteristics (symptoms, diagnosis, complications), laboratory data (e.g., lipid profile, FBS, CK-MB), imaging and procedure data (echocardiography, CAG, PCI), treatment details, and outcomes (discharge, mortality).

### 4.7 A pilot test

A pilot test on 10% of records assessed checklist clarity and consistency. Revisions were made accordingly before full data collection.

### 4.8 Validity and Reliability

Face and content validity were confirmed by experts in cardiology and epidemiology. The checklist, adapted from validated tools, showed good inter-rater reliability (κ = 0.70).

### 4.9 Statistical Analysis Data were analyzed using SPSS (version 2024)

Data were analyzed using SPSS v.2024. Descriptive statistics, chi-square tests, and one-way ANOVA assessed distributions and group differences. Univariate logistic regression identified predictors of in-hospital and 5-year mortality, reporting ORs, CIs, and p-values

### 4.10 Study Variables Dependent variable

The outcome variable was mortality (in-hospital and 5-year); predictors included sociodemographic, clinical, risk, and treatment variables.

### 4.11 Ethical Consideration

The study received ethical approval from Al-Razi University and access permission for patient records was obtained from the Military Cardiac Center management.

## 5.0 Result

### 5.1 Age Group Distribution

The mean age of patients admitted to the CCU was 53.6 ± 13.6 years. The age group of 18–30 years was represented as 7%, from 31-40 years 8%, from 41 – 50 years 26%, from 51-60 years 34%, from 61-70 years 16%, and 9% represents> 70 years.

### 5.2. Distribution of Cases by Final Diagnosis and Sex (Table 5.1)

that acute coronary syndrome (60.9%)—mainly STEMI (52%)—was the most common diagnosis, especially in males. Heart failure (13%) and rheumatic valvular disease (9%) were more common in females (pü<ü0.001 and pü=ü0.036). Less frequent conditions included cardiomyopathy (4%), cardiogenic shock (5%), and others (2.2%). The data emphasize the high burden of ischemic heart disease in males and structural heart disease in females, highlighting the need for gender-sensitive care in Yemen

**Table 5.1:** Distribution of cases by final diagnosis of the admission event and Sex.

| Final diagnosis of the admission event | Male no (%) | Female no (%) | Total | P.value |
| --- | --- | --- | --- | --- |
| 1. ACS | 241 (64%) | 31 (44%) | 272 (60.9%) | **0.002** |
| 2. ACS STEMI | 131 (35%) | 12 (17%) | 143 (52.3) | **0.002** |
| 3. ACS NSTEMI | 52 (14%) | 8 (11%) | 60 (22) | 0.374 |
| 4. ACS SA | 5 (1%) | 0 (0%) | 5 (1.8%) | 0.424 |
| 5. ACS UA | 53 (14%) | 11 (16%) | 64 (23.5%) | 0.421 |
| 6. ADHF | 47 (13%) | 10 (14%) | 57 (13%) | 0.402 |
| 7. Arrhythmia | 4 (1%) | 1 (1%) | 5 (1%) | 0.576 |
| 8. Cardiogenic Shock | 21 (6%) | 2 (3%) | 23 (5%) | 0.270 |
| 9. Congenital Heart Disease | 11 (3%) | 6 (9%) | 17 (4%) | **0.036** |
| 10. COPD | 3 (1%) | 1 (1%) | 4 (1%) | 0.496 |
| 11. DCM | 17 (5%) | 1 (1%) | 18 (4%) | 0.195 |
| 12. Hypertensive Emergency | 1(0%) | 1(1%) | 2(0%) | 0.290 |
| 13. LV THROMBUS | 2 (1%) | 0 (0%) | 2 (0%) | 0.710 |
| 14. P.Embolism | 2 (0%) | 0 (0%) | 2 (0%) | 0.710 |
| 15. P.HTN | 1 (0%) | 1 (1%) | 2 (0%) | 0.290 |
| 16. Pericarditis | 2 (1%) | 0 (0%) | 2 (0%) | 0.710 |
| 17. VHD | 24 (6%) | 16 (23%) | 40 (9%) | **0.000** |
| **TOTAL** | **376 (100%)** | **70 (100%)** | **446 (100%)** |  |

### 5.3. Frequency of CVD risk factors among cases (Table 5.2)

**As shown in Table 5.2**, khat chewing (76%) and smoking (61%) were the most frequent risk factors, followed by hypertension (41%) and diabetes (31%). About 40% had three or more risk factors. Men reported more khat, smoking, and shammah users, while women had higher rates of hypertension, diabetes, and dyslipidemia, highlighting the need for gender-specific prevention strategies.

**Table 5.2:** Frequency of CVD Risk Factors among Cases.

| Risk Factor | Male |  |  |  | Female |  |  |  | Total |
| --- | --- | --- | --- | --- | --- | --- | --- | --- | --- |
|  | Yes | % | NO | % | Yes | % | No | % | Yes % |
| DM | 114 | 30% | 262 | 70% | 24 | 34% | 46 | 66% | 31% (138) |
| Hypertension | 150 | 40% | 226 | 60% | 32 | 46% | 38 | 54% | 41% (182) |
| Smoking | 242 | 65% | 131 | 35% | 31 | 44% | 39 | 56% | 61% (273) |
| Khat chewing | 304 | 81% | 72 | 19% | 35 | 50% | 35 | 50% | 76% (339) |
| Shammah users | 64 | 17% | 312 | 83% | 3 | 4% | 67 | 96% | 15% (67) |
| Dyslipidemia | 16 | 4% | 360 | 96% | 6 | 9% | 64 | 91% | 4.9% (22) |
| Daily smoker | 154 | 41% | 222 | 59% | 16 | 23% | 54 | 77% | 38% (170) |

### 5.4 Baseline Demographic and Clinical Characteristics (Table 5.3)

**As shown in Table 5.3**, presents data on 446 CCU patients at the Military Cardiac Center in Sana’a (2019), categorized into three age groups: ≤50 years (41%), 51–70 years (50%), and >70 years (9%), highlighting the predominance of middle-aged patients.

**Table 5.3:**
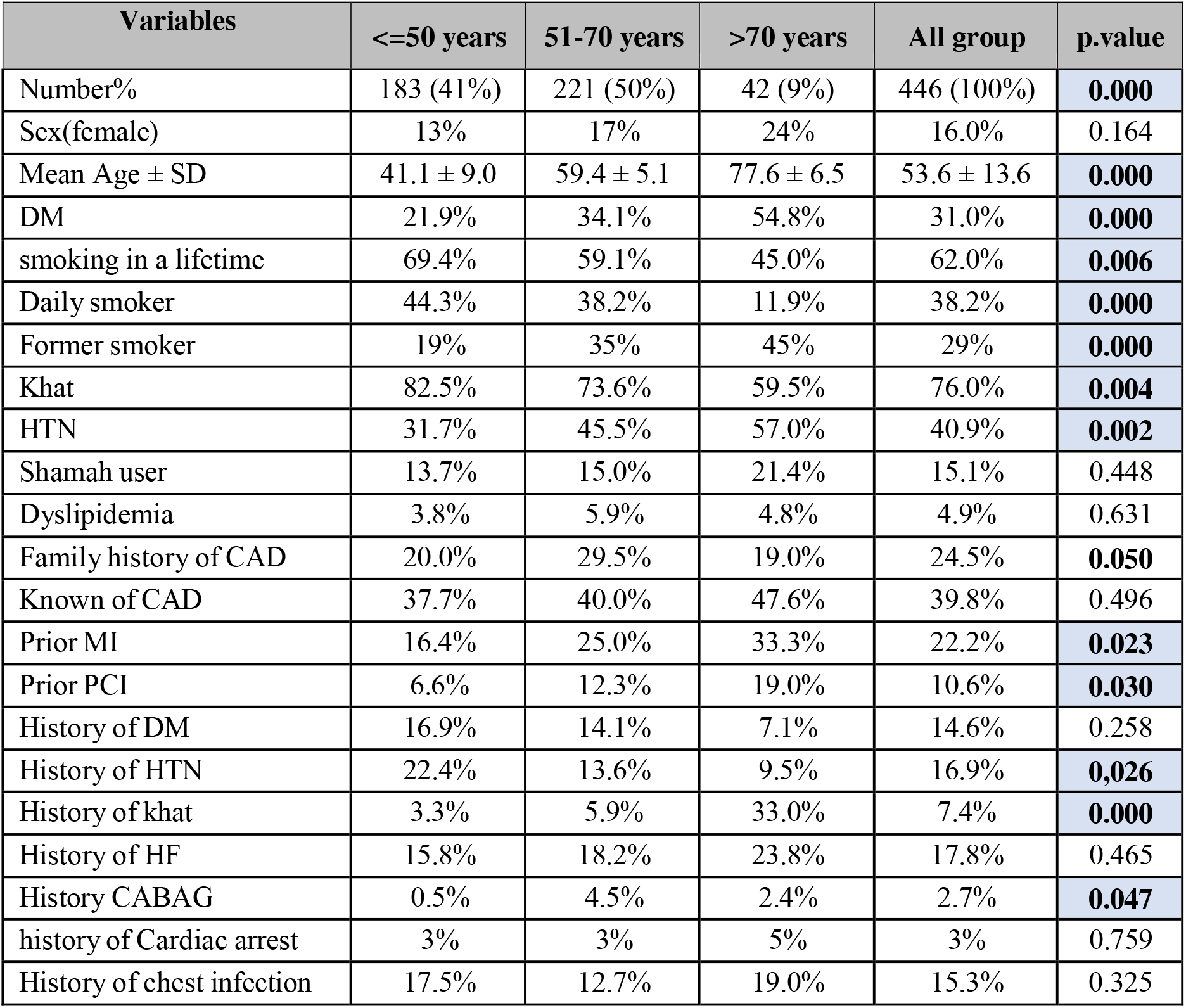
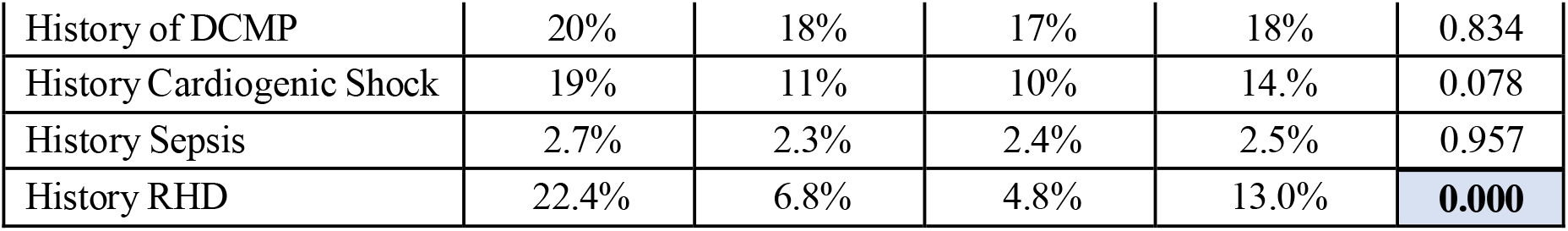
Baseline Demographic and Clinical Characteristics.

| Variables | $\leq 50$ years | 51-70 years | $> 70$ years | All group | p.value |
| --- | --- | --- | --- | --- | --- |
| Number% | 183 (41%) | 221 (50%) | 42 (9%) | 446 (100%) | **0.000** |
| Sex(female) | 13% | 17% | 24% | 16.0% | 0.164 |
| Mean Age ± SD | 41.1 ± 9.0 | 59.4 ± 5.1 | 77.6 ± 6.5 | 53.6 ± 13.6 | **0.000** |
| DM | 21.9% | 34.1% | 54.8% | 31.0% | **0.000** |
| smoking in a lifetime | 69.4% | 59.1% | 45.0% | 62.0% | **0.006** |
| Daily smoker | 44.3% | 38.2% | 11.9% | 38.2% | **0.000** |
| Former smoker | 19% | 35% | 45% | 29% | **0.000** |
| Khat | 82.5% | 73.6% | 59.5% | 76.0% | **0.004** |
| HTN | 31.7% | 45.5% | 57.0% | 40.9% | **0.002** |
| Shamah user | 13.7% | 15.0% | 21.4% | 15.1% | 0.448 |
| Dyslipidemia | 3.8% | 5.9% | 4.8% | 4.9% | 0.631 |
| Family history of CAD | 20.0% | 29.5% | 19.0% | 24.5% | **0.050** |
| Known of CAD | 37.7% | 40.0% | 47.6% | 39.8% | 0.496 |
| Prior MI | 16.4% | 25.0% | 33.3% | 22.2% | **0.023** |
| Prior PCI | 6.6% | 12.3% | 19.0% | 10.6% | **0.030** |
| History of DM | 16.9% | 14.1% | 7.1% | 14.6% | 0.258 |
| History of HTN | 22.4% | 13.6% | 9.5% | 16.9% | **0.026** |
| History of khat | 3.3% | 5.9% | 33.0% | 7.4% | **0.000** |
| History of HF | 15.8% | 18.2% | 23.8% | 17.8% | 0.465 |
| History CABAG | 0.5% | 4.5% | 2.4% | 2.7% | **0.047** |
| history of Cardiac arrest | 3% | 3% | 5% | 3% | 0.759 |
| History of chest infection | 17.5% | 12.7% | 19.0% | 15.3% | 0.325 |
| History of DCMP | 20% | 18% | 17% | 18% | 0.834 |
| History Cardiogenic Shock | 19% | 11% | 10% | 14.4% | 0.078 |
| History Sepsis | 2.7% | 2.3% | 2.4% | 2.5% | 0.957 |
| History RHD | 22.4% | 6.8% | 4.8% | 13.0% | **0.000** |

#### Age and Sex Distribution

The mean patient age was 53.6 ± 13.6 years (p < 0.001), with most patients in the 51–70 age group. Female representation rose with age—13% in ≤50 years, 17% in 51–70 years, and 24% in >70 years. While the overall sex distribution was not statistically significant (p = 0.164), the age-related increase in female proportion was notable

#### Cardiovascular Risk Factors by Age

Diabetes and hypertension increased significantly with age (pü<ü0.001 and pü=ü0.002), while smoking and khat use were more common among younger patients (pü<ü0.01). For instance, khat use was 82.5% in those ≤50 years versus 59.5% in those >70, reflecting generational behavioral differences.

#### Clinical History and Comorbidities

Older patients had higher rates of diabetes, hypertension, dyslipidemia, shammah use, and past cardiac interventions (MI, PCI, CABG) (p**∬**<**∬**0.05). In contrast, smoking and khat use were more common in younger individuals. Rheumatic heart disease (RHD) was more prevalent in younger patients, likely due to early-life exposures. A family history of CAD was notably reported in the middle-aged group, aligning with regional trends in the Gulf States **(5**).

### 5.5 Clinical and Laboratory Characteristics by Age Group (Table 5.4)

**As shown in Table 5.4** the distribution of clinical signs, laboratory findings, and presenting symptoms varied significantly across age groups, revealing several important trends.

**Table 5.4:** Baseline demographic and clinical characteristics.

| Variables | $\leq 50$ years | 51-70 years | $>70$ years | All group | p.value |
| --- | --- | --- | --- | --- | --- |
| DBP (mmHg) (mean SD) | 68 ± 16 | 69 ± 14 | 70 ± 17.6 | 69.1 ± 15 | 0.925 |
| SBP (mmHg) (mean SD) | 111 ± 22 | 113 ± 25 | 125 ± 29 | 113 ± 25 | **0.005** |
| Positive cardiac enzyme | 72% | 67% | 50% | 68% | **0.024** |
| HR (BPM) (mean SD) | 92 ± 18 | 88 ± 20 | 85 ± 26 | 89 ± 20 | 0.071 |
| CK (mean ± SD) | 530 ± 780 | 440 ± 618 | 391 ± 580 | 473 ± 688 | 0.349 |
| CKMP (mean ± SD) | 78 ± 107 | 70 ± 82 | 87 ± 139 | 75 ± 99 | 0.517 |
| FBS mmol/l (mean ± SD) | 138 ± 72 | 142 ± 72 | 146 ± 58 | 140 ± 71 | 0.181 |
| RBS mmol/l (mean ± SD) | 142 ± 71 | 167 ± 106 | 180 ± 80 | 158 ± 92 | **0.006** |
| HBA1C | 7.7 ± 3 | 8.0 ± 2.7 | 8.2 ± 2.7 | 8.1 ± 2.8 | 0.810 |
| TC mg/dl/l (mean ± SD) | 148 ± 41 | 144 ± 42 | 149 ± 44 | 146 ± 42 | 0.555 |
| TG mg/dl (mean ± SD) | 169 ± 100 | 160 ± 84 | 143 ± 74 | 162 ± 90 | 0.332 |
| LDL mg/dl (mean ± SD) | 90.8 ± 34 | 85.6 ± 34 | 93 ± 31 | 88.3 ± 34 | 0.293 |
| HDL mg/dl (mean ± SD) | 33 ± 8.8 | 36.3 ± 27 | 35.8 ± 18 | 35.0 ± 21 | 0.407 |
| Chest pain (any) | 55.7% (102) | 72.4% (160) | 69.0% (29) | 65.2% (291) | 0.002 |
| Typical chest pain | 36.6% (67) | 50.7% (112) | 57.1% (24) | 45.5% (203) | **0.005** |
| Atypical chest pain | 16.4% (30) | 29.9% (66) | 45.2% (19) | 25.8% (115) | **<0.001** |
| Dyspnea | 38.3% (70) | 56.1% (124) | 66.7% (28) | 49.8% (222) | **<0.001** |
| ST-elevation MI | 43.7% (80) | 26.7% (59) | 9.5% (4) | 52.6% (143/272) | **<0.001** |
| NST-elevation MI | 5.5% (10) | 18.1% (40) | 23.8% (10) | 22.1% (60/272) | **<0.001** |
| Unstable angina | 7.7% (14) | 18.1% (40) | 23.8% (10) | 23.5% (64/272) | **0.002** |
| Stable angina | 1.6% (3) | 0.9% (2) | 0.0% (0) | 1.8% (5/272) | 0.603 |

#### Blood Pressure and Blood Sugar Trends-

Patients over 70 years of age had the highest systolic blood pressure (SBP), diastolic blood pressure (DBP), and CK-MB enzyme levels, with SBP differences reaching statistical significance (p = 0.005). In contrast, younger patients (≤50 years) had higher levels of total CK and heart rate (HR), though these differences were not statistically significant. The rate of positive cardiac enzyme findings was notably higher among younger patients (72%) compared to older groups (p = 0.024),

#### Glycemic and Lipid Profiles

Random blood sugar (RBS) levels increased significantly with age, with patients >70 years showing the highest mean values (p = 0.006). Fasting blood sugar (FBS), HbA1c, total cholesterol (TC), low-density lipoprotein (LDL), and high-density lipoprotein (HDL) levels were also higher in older age groups, although most differences did not reach statistical significance. These findings suggest that older patients were more likely to present with metabolic comorbidities such as diabetes and dyslipidemia.

#### Chest Pain Presentation and Dyspnea

Chest pain was reported in 65.2% of patients, with significant differences across age groups (55.7%, 72.4%, and 69.0% among patients aged ≤50, 51–70, and >70 years, respectively; *p* = 0.002). Typical chest pain increased with age (36.6%, 50.7%, and 57.1%; *p* = 0.005), as did atypical chest pain (16.4%, 29.9%, and 45.2%; *p* < 0.001). Dyspnea was also more frequent with increasing age, affecting 38.3%, 56.1%, and 66.7% of patients, respectively (*p* < 0.001).

#### Acute Coronary Syndrome Classification

STEMI was most frequent among younger patients, decreasing from 43.7% among those aged ≤50 years to 26.7% among those aged 51–70 years and 9.5% among those aged >70 years (*p* < 0.001). Conversely, NSTEMI increased across these age groups (5.5%, 18.1%, and 23.8%; *p* < 0.001), as did unstable angina (7.7%, 18.1%, and 23.8%; *p* = 0.002). Stable angina was uncommon and did not differ significantly across age groups (1.6%, 0.9%, and 0.0%; *p* = 0.603)

### 5.6 Medical Therapy and Outcomes (Table 5.5)

**As shown in Table 5.5**, Medical therapy varied by age. Patients >70 years received more beta-blockers (62%), clopidogrel (76%), statins (74%), ACE inhibitors (55%), and antibiotics (88%) (p**∬**<**∬**0.05), indicating more intensive care. Younger patients (≤50 years) were more likely to receive pantoprazole (41%), metoprolol (12%), and digoxin (7%). Common therapies overall included aspirin (90%), clopidogrel (74%), statins (63%), antibiotics (70%), ACE inhibitors (35%), and diuretics (43%). Use of dopamine, thrombolytics, and GP IIb/IIIa inhibitors was limited. These trends suggest age-specific pharmacologic management tailored to cardiovascular risk and clinical status.

**Table 5.5:**
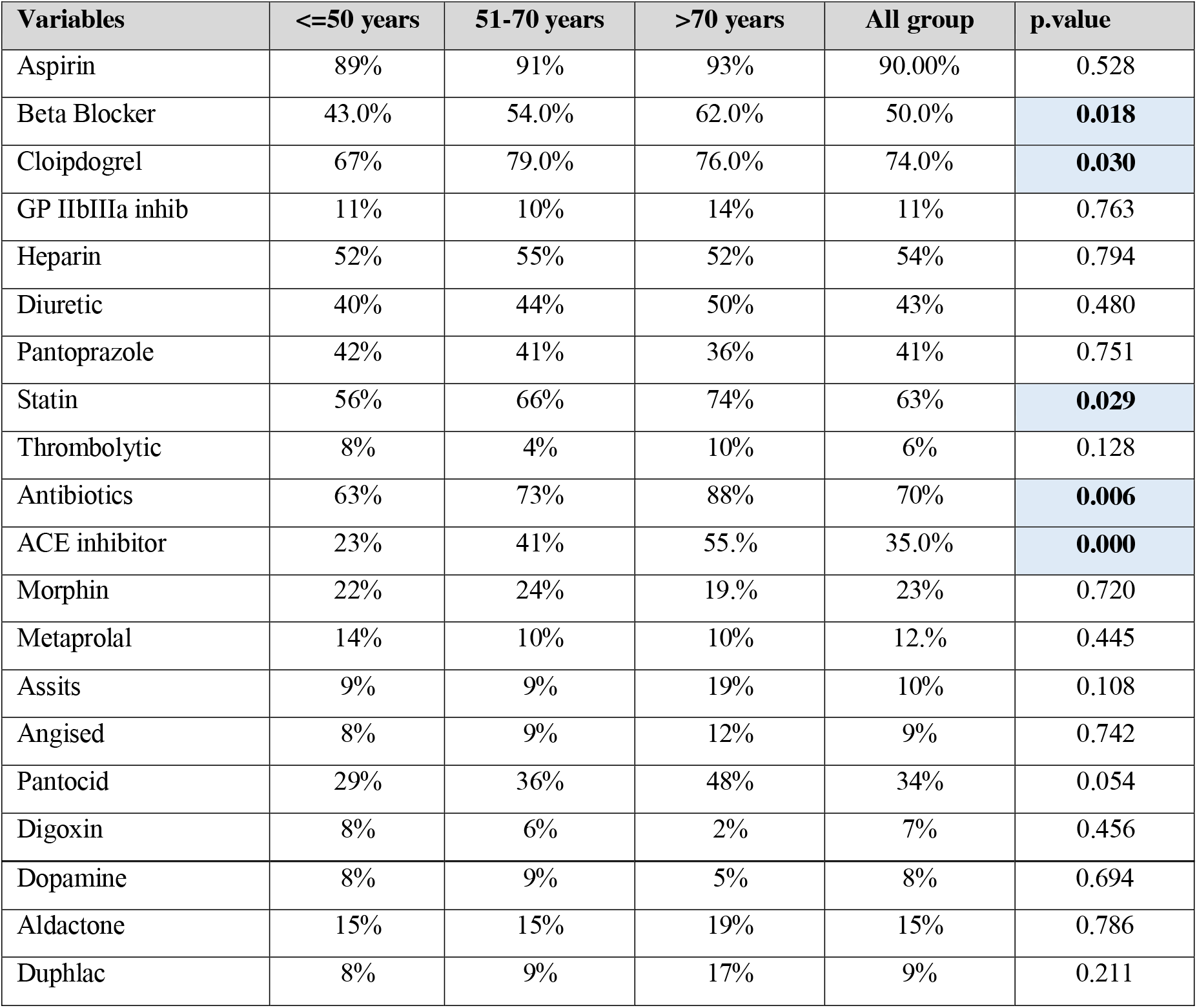
Medical therapy.

| Variables | <=50 years | 51-70 years | >70 years | All group | p.value |
| --- | --- | --- | --- | --- | --- |
| Aspirin | 89% | 91% | 93% | 90.00% | 0.528 |
| Beta Blocker | 43.0% | 54.0% | 62.0% | 50.0% | **0.018** |
| Clopidogrel | 67% | 79.0% | 76.0% | 74.0% | **0.030** |
| GP IIb/IIIa inhib | 11% | 10% | 14% | 11% | 0.763 |
| Heparin | 52% | 55% | 52% | 54% | 0.794 |
| Diuretic | 40% | 44% | 50% | 43% | 0.480 |
| Pantoprazole | 42% | 41% | 36% | 41% | 0.751 |
| Statin | 56% | 66% | 74% | 63% | **0.029** |
| Thrombolytic | 8% | 4% | 10% | 6% | 0.128 |
| Antibiotics | 63% | 73% | 88% | 70% | **0.006** |
| ACE inhibitor | 23% | 41% | 55.0% | 35.0% | **0.000** |
| Morphin | 22% | 24% | 19.0% | 23% | 0.720 |
| Metoprolol | 14% | 10% | 10% | 12.0% | 0.445 |
| Assits | 9% | 9% | 19% | 10% | 0.108 |
| Angised | 8% | 9% | 12% | 9% | 0.742 |
| Pantocid | 29% | 36% | 48% | 34% | 0.054 |
| Digoxin | 8% | 6% | 2% | 7% | 0.456 |
| Dopamine | 8% | 9% | 5% | 8% | 0.694 |
| Aldactone | 15% | 15% | 19% | 15% | 0.786 |
| Duphac | 8% | 9% | 17% | 9% | 0.211 |

### 5.7 Clinical Outcomes (Table 5.6)

**As shown in Table 5.6**, Complications occurred in 40.8% of patients, with nosocomial infections (25%, p = 0.008), septicemia (7%, p = 0.000), and chest infections (7%) being the most common. Other complications included mural thrombus (4%), bleeding, emergency surgery, renal impairment, and conduction block (1–3%). Khat chewers had significantly more complications (31.1%) than non-chewers (9.4%), consistent with reports linking khat to increased stroke and mortality risk **(7)**.In-hospital mortality was higher in patients >70 years (17%) compared to ≤50 years (13%). Five-year mortality also followed this trend (21% vs. 20%), in line with regional findings **(5)**.

**Table 5.6:** Outcome and Mortality.

| Variables | ≤50 years | 51-70 years | >70 years | All group | p.value |
| --- | --- | --- | --- | --- | --- |
| Infection | 32% | 22% | 12% | 25% | **0.008** |
| Cardiac valve injury | 2% | 0% | 0% | 1% | 0.368 |
| L V thrombus | 3% | 3% | 0% | 3% | 0.499 |
| Mural thrombus | 6% | 4% | 0% | 4% | 0.218 |
| Blood transfusion | 3% | 2% | 2% | 3% | 0.815 |
| Urgent cardiac surgery | 4% | 3% | 2% | 3% | 0.874 |
| Renal Impermeant | 1% | 2% | 5% | 2% | 0.136 |
| Chest infection | 6% | 8% | 2% | 7% | 0.416 |
| Sepsis | 7% | 5% | 14% | 7% | **0.000** |
| P. Oedema | 0% | 1% | 2% | 1% | 0.200 |
| Conduction block | 2% | 1% | 0% | 2% | 0.555 |
| Cardiac arrest | 4% | 5% | 7% | 4% | 0.646 |
| Diarrhea | 1% | 1% | 0% | 1% | 0.777 |
| Warfarin Toxicity | 1% | 1% | 2% | 1% | 0.103 |
| In hospital mortality | 13% | 15% | 17% | 14% | 0.791 |
| Mortality during 5 years | 20% | 18% | 21% | 19% | 0.809 |

### 5.8 Distribution of Overall Outcomes by Final Diagnosis (Table 5.7)

**As shown in Table 5.7** The overall in-hospital mortality rate was 14.3% (64/446). Acute coronary syndrome (ACS) was the primary cause, accounting for 26 deaths (5.8%, p = 0.000), with STEMI contributing 19 (4.3%) and NSTEMI 5 (1.1%). Unstable angina was significantly associated with mortality (p = 0.001). Acute decompensated heart failure (ADHF) caused 15 deaths (3.4%, p = 0.008), and cardiogenic shock 7 (1.6%, p = 0.033). Valve heart disease accounted for another 7 deaths (1.6%). Less common fatal diagnoses included dilated cardiomyopathy (DCMP), pulmonary hypertension (P.HTN, p = 0.020), hypertensive emergencies, and pericarditis. These data emphasize the strong link between ischemic and decompensated cardiac conditions and mortality in this population.

**Table 5.7:** Distribution of the overall outcome of the patients by their diagnosis.

| Final diagnosis | Alive | % | Dead | % | TOTAL | P.value |
| --- | --- | --- | --- | --- | --- | --- |
| ACS | 246 | 55.1% | 26 | 5.8% | 272 | **0.000** |
| ACS (STEMI) | 124 | 27.8% | 19 | 4.3% | 143 | 0.388 |
| ACS (NSTEMI) | 55 | 12.3% | 5 | 1.1% | 60 | 0.105 |
| ACS (Unstable) | 62 | 13.9% | 2 | 0.4% | 64 | **0.001** |
| ACS SA | 5 | 1.1% | 0 | 0.0% | 5 | 0.105 |
| ADHF | 42 | 9.4% | 15 | 3.4% | 57 | **0.008** |
| Arrhythmia | 5 | 1.1% | 0 | 0.0% | 5 | 0.459 |
| Cardiogenic Shock | 16 | 3.6% | 7 | 1.6% | 23 | **0.033** |
| Congenital heart disease | 17 | 3.8 | 0 | 0.0% | 17 | 0.068 |
| COPD | 4 | 0.9% | 0 | 0.0% | 4 | 0.537 |
| DCMP | 14 | 3.1% | 4 | 0.9% | 18 | 0.250 |
| Hypertensive Emergency | 1 | 0.2% | 1 | 0.4% | 2 | 0.267 |
| LV THROMBUS | 1 | 0.2% | 1 | 0.2% | 2 | 0.267 |
| P.Embolism | 2 | 0.4% | 0 | 0.0% | 2 | 0.733 |
| P.HTN | 0 | 0.0% | 2 | 0.4% | 2 | **0.020** |
| Pericarditis | 1 | 0.2% | 1 | 0.2% | 2 | 0.267 |
| VHD | 33 | 7.4% | 7 | 1.6% | 40 | 0.345 |
| **TOTAL** | **382** | **5.8%** | **64** | **14.3%** | **446** |  |

### 5.9 Risk Factors Associated with Cardiovascular Diagnoses (Table 5.8)

**As shown in Table 5.8**,, acute coronary syndrome (ACS) was significantly associated with delayed smoking (p = 0.000) and diabetes (p = 0.002). STEMI showed associations with smoking (p = 0.004), hypertension (p = 0.012), diabetes (p = 0.015), khat chewing (p = 0.009), and delayed smoking (p = 0.000). NSTEMI was linked to shammah users (p = 0.046), while unstable angina was related to smoking (p = 0.005) and khat (p = 0.014). Heart failure subtypes also showed strong associations: ADHF with smoking (p = 0.017), diabetes (p = 0.000), and hypertension (p = 0.002), while rheumatic heart disease was linked to smoking (p = 0.007), hypertension (p = 0.001), diabetes (p = 0.000), and delayed smoking (p = 0.009). These findings underscore the impact of modifiable risk factors on cardiovascular outcomes.

**Table 5.8:** Risk factors associated with CVD patient diagnosis.

| final Diagnosis | ACS | STEMI | NSTEMI | U.A | ADHF | C, shock | DCMP | RHD |
| --- | --- | --- | --- | --- | --- | --- | --- | --- |
| Smoking | 174 | 102 | 40 | 30 | 43 | 15 | 11 | 17 |
| P.value | 0.180 | **0.004** | 0.263 | **0.005** | **0.017** | 0.469 | 0.556 | **0.007** |
| HTN | 105 | 47 | 26 | 31 | 34 | 12 | 8 | 7 |
| P.value | 0.139 | **0.012** | 0.385 | 0.115 | **0.002** | 0.178 | 0.469 | **0.001** |
| DM | 70 | 34 | 138 | 16 | 34 | 7 | 9 | 2 |
| P.value | **0.002** | **0.015** | 0.502 | 0.167 | **0.000** | 0.580 | 0.067 | **0.000** |
| Khat user | 213 | 119 | 49 | 41 | 43 | 17 | 15 | 30 |
| P.value | 0.096 | **0.009** | 0.174 | **0.014** | 0.514 | 0.488 | 0.337 | 0.504 |
| Shammah users | 45 | 67 | 14 | 24 | 10 | 3 | 3 | 3 |
| P.value | 0.161 | 0.382 | **0.046** | 0.515 | 0.344 | 0.537 | 0.524 | 0.118 |
| Dely smoking | 125 | 76 | 23 | 8 | 16 | 5 | 4 | 8 |
| P.value | **0.000** | **0.000** | 0.539 | 0.347 | 0.062 | 0.072 | 0.119 | **0.009** |

### 5.10 Predictors of In Hospital Mortality – Univariate Binary Logistic Regression (Table 5.9)

**As shown in Table 5.9**, univariate binary logistic regression identified several variables significantly associated with in-hospital mortality (p < 0.05)..

**Table 5.9:** Univariate Binary Logistic Regression Analysis for In-Hospital Mortality.

| Variable | In hospital mortality |  |  |
| --- | --- | --- | --- |
|  | OR | 95% C. L | p.value |
| Gender | 1.6 | 0.3 – 3.7 | 0.260 |
| Age | 1.4 | 0.4 – 4.6 | 0.450 |
| Hypertension | 1.6 | 0.9 – 2.6 | 0.108 |
| Khat | 1.4 | 0.8 – 2.5 | 0.250 |
| D.M | 2.6 | 1.5 - 4.5 | **0.000** |
| Smoking | 1.19 | 0.7 – 2.1 | 0.531 |
| History of CAD | 1.0 | 0.5 – 1.9 | 0.910 |
| History of RHD | 2.4 | 1.2 – 4.7 | **0.009** |
| History of Cardiogenic Shock | 11.3 | 6.1 – 20.9 | **0.000** |
| History of Cardiac Arrest | 5.5 | 1.8 – 17 | **0.003** |
| History of DCMP | 3.6 | 2.0 – 6.5 | **0.000** |
| F.History of DM | 1.6 | 0.82 -3.2 | 0.163 |
| History of a former smoker | 2.1 | 1.2 – 3.6 | **0.007** |
| History of Heart failure | 9.2 | 5.1 – 16.6 | **0.000** |
| History of Renal Impairment | 3.5 | 1.5 – 8.3 | **0.004** |
| History of post-SK | 4.5 | 1.2 – 16.4 | **0.023** |
| History of Chronic Lung Disease | 8.0 | 2.1 – 30.6 | **0.002** |
| History of Oedema | 3.2 | 1.5 – 6.36 | **0.001** |
| History of P.Effusion | 10.7 | 2.4 – 45.9 | **0.001** |
| Presentation of Sepsis | 31.1 | 6.5 - 147 | **0.000** |
| Presentation of Chest Infection | 2.6 | 1.4 – 4.8 | **0.003** |
| Presentation of Septic Shock | 8.0 | 2.1 – 30.7 | **0.002** |
| Presentations CPR in ER | 4.1 | 1.5 – 10.9 | **0.005** |
| CCU CPR | 1.1 | 0.1 – 3.0 | 0.899 |
| Presentation of Shocked liver | 13.2 | 1.1 - 148 | **0.036** |
| Presentation of Dyspnea | 6.9 | 3.4 – 13.9 | **0.000** |
| Presentation of Dyspnea Class II | 3.5 | 1.8 – 6.9 | **0.000** |
| Presentation of Dyspnea Class III | 2.7 | 1.4 – 5.1 | **0.002** |
| Presentation of Dyspnea Class IV | 2.2 | 1.1 – 4.5 | **0.022** |
| Positive Cardiac enzyme | 3.9 | 1.8 – 8.4 | **0.001** |
| CKMP | 3.3 | 1.4 – 7.8 | **0.004** |
| Aldactone | 2.1 | 1.1 - 3.9 | **0.025** |
| Duphlac | 3.6 | 1.8 – 7.5 | **0.000** |
| ACS | 2.6 | 1.5 – 4.5 | **0.000** |
| ACS (STEMI) | 0.9 | 0.4 – 1.6 | 0.660 |
| ACS (NSTEMI) | 0.5 | 0.19 – 1.3 | 0.160 |
| ACS US | 0.2 | 0.4 – 0.7 | 0.014 |
| VHD | 1.3 | 0.5 – 3.0 | 0.552 |
| ADHF | 2.5 | 1.3 – 4.8 | **0.007** |
| Cardiogenic shock | 2.8 | 1.1 – 7.1 | **0.030** |
| DCMP | 1.8 | 0.6 – 5.5 | 0.337 |
| Chest pain | 1.9 | 1.1 – 3.3 | **0.014** |
| Typical chest pain | 1.5 | 0.9 – 2.7 | 0.098 |
| Atypical chest pain | 2.3 | 1.1 – 4.9 | **0.024** |
| Age group | 1.3 | 0.5 – 3.3 | 0.548 |
| 51-70 years | 1.1 | 0.6 – 1.8 | 0.728 |
| >70 years | 1.2 | 0.5 – 2.8 | 0.653 |
| Complication of Infection | 2.5 | 1.5 – 4.5 | **0.001** |
| Complications of Cardiac arrest | 74 | 16.7 - 330 | **0.000** |
| Complication of P.odema | 18.7 | 1.9 - 183 | **0.012** |
| Complication of Shocked liver | 9.7 | 2.8 – 35.7 | **0.001** |
| Complication of Sepsis | 5.8 | 2.7 – 12.6 | **0.000** |
| Complications of Cardiac Valve Injury | 18.7 | 1.9 – 183 | **0.012** |

#### Clinical history

Key predictors included diabetes mellitus (OR**∬**2.6), rheumatic heart disease (OR**∬**2.4), dilated cardiomyopathy (OR**∬**3.6), heart failure (OR**∬**9.2), renal impairment (OR**∬**3.5), chronic lung disease (OR**∬**8.0), oedema (OR**∬**3.2), and former smoking (OR**∬**2.1).

#### Presentation features

Strong predictors included cardiogenic shock (OR**∬**11.3), cardiac arrest (OR**∬**5.5), dyspnea (OR**∬**6.9), positive cardiac enzymes (OR**∬**3.9), and elevated CK-MB (OR**∬**3.3).

#### Diagnostic categories

Mortality was associated with acute coronary syndrome (OR**∬**2.6), cardiogenic shock (OR**∬**2.8), and acute decompensated heart failure (OR**∬**2.5), while unstable angina was protective (OR**∬**0.2).

#### In hospital complications

The strongest predictors were complications of cardiac arrest (OR**∬**74), sepsis (OR**∬**31.1), pulmonary oedema (OR**∬**18.7), shocked liver (OR**∬**9.7), and infection (OR**∬**2.5).

These findings emphasize the importance of early recognition and management of high-risk factors to improve hospital outcomes.

## 5.11 Predictors of In Hospital and 5 Year Mortality (Table 5.10)

**As shown in Table 5.10**, univariate binary logistic regression identified several variables significantly associated with both in-hospital and 5-year mortality. Key findings include:

**Table 5.10:** Univariate Binary Logistic Regression for In-Hospital and within 5-Year Mortality.

| Variable | In hospital mortality |  |  | Mortality within 5 years |  |  |
| --- | --- | --- | --- | --- | --- | --- |
|  | OR | 95% C. L | p.value | OR | 95% C. L | p.value |
| Khat | 0.71 | 0.4 – 1.2 | 0.009 | 2.0 | 1.1 – 3.2 | 0.009 |
| Clopidogrel | 2.5 | 1.5 – 4.0 | 0.001 | 2.5 | 1.5 – 4.0 | 0.001 |
| Beta-blocker | 2.3 | 1.3 - 4.1 | 0.003 | 2.1 | 1.3 – 3.6 | 0.002 |
| Metoprolol | 3.0 | 0.9 - 10 | 0.067 | 3.2 | 1.1 – 9.2 | 0.029 |
| Statin | 1.8 | 1.0 - 3.1 | 0.026 | 2.4 | 1.5 – 3.9 | 0.000 |
| Elective PCI | 2.6 | 1,1 – 5.9 | 0.021 | 2.9 | 1.0 – 8.3 | 0.004 |
| Lt Catheterization | 4.9 | 2.6 - 9.2 | 0.000 | 3.5 | 2.1 – 5.7 | 0.000 |
| PPCI | 0.88 | 0.2 – 2.9 | 0.77 | 1.2 | 0.3 – 4.2 | 0.775 |
| ACS | 2.6 | 1.5 – 4.5 | 0.000 | 4.0 | 2.4 – 6.6 | 0.000 |
| ACS US | **0.2** | 0.4 – 0.7 | **0.014** | **8.7** | 2.3 – 36 | **0.002** |
| Chest pain | **1.9** | 1.1 – 3.3 | **0.014** | **2.6** | 1.6 1 4.2 | **0.000** |
| Typical chest pain | 1.5 | 0.9 – 2.7 | 0.098 | **2.3** | 1.4 – 4.0 | **0.001** |
| Atypical chest pain | **2.3** | 1.1 – 4.9 | **0.024** | **2.0** | 1.0 -3.7 | **0.027** |
| Complications of cardiac arrest | **74** | 16.7 - 330 | **0.000** | 0.01 | 0.01 – 0.7 | 0.000 |

**Pharmacologic and procedural interventions**, Clopidogrel (OR**∬**2.5), beta-blockers (OR**∬**2.3 and 2.1), and statins (OR**∬**1.8 and 2.4) were significantly associated with increased mortality. Elective PCI (OR**∬**2.6 and 2.9) and left heart catheterization (OR**∬**4.9 and 3.5) predicted mortality in both periods.

### Symptoms and clinical presentation

Chest pain (OR**∬**1.9 and 2.6) and atypical chest pain (OR**∬**2.3 and 2.0) predicted higher mortality. Unstable angina was protective during hospitalization (OR**∬**0.2) but linked to higher long-term risk (OR**∬**8.7).

### Diagnosis and outcomes

ACS increased mortality in both periods (OR**∬**2.6 and 4.0). Cardiac arrest complications were the strongest in-hospital predictor (OR**∬**74).

### Procedural outcomes

Even though PPCI was not statistically linked to a lower death risk (OR**∬**0.88 and 1.2), its early use might still help patients and lead to better recovery.

These results show that different factors affect short-term and long-term mortality, and emphasize the need for personalized treatment strategies at each stage of care.

## 6.0 Discussion

### 6.1. Age Group Distribution

The current study showed that only 7% of patients were aged 18–30 years, a figure higher than reports from Germany (1.7%) and New York (1.8%) but close to Bangladesh (9%)***(8-10)***. This may reflect early exposure to modifiable risk factors like khat chewing, smoking, and a sedentary lifestyle.

The most affected group was aged 51–60 years (34%), which contrasts with findings in Europe, where older patients predominate. For example, 30.3% of patients in the Netherlands were over 75, and 55.5% in Denmark were over 70 ***(11, 12)***. In Yemen, only 9% were over 70.

Notably, 41% of Yemeni patients were under 50, compared to only 9% in Denmark and 8% in the Netherlands. This suggests early onset of cardiovascular disease in Yemen, likely due to poor access to preventive care and social determinants such as poverty and stress. These disparities underscore the need for youth-focused cardiovascular screening and public health interventions in low-resource settings.

### 6.2. Distribution of Cases by Final Diagnosis and Sex

STEMI was more common in males, whereas RHD and congenital heart disease were more prevalent in females, aligning with global patterns. Male dominance (84%) in this study was notably higher than in Egypt (74%), Saudi Arabia (72.3%), and North America (50–63%) ***(13-15)*** This skewed ratio may be due to the military hospital setting and barriers limiting female access to care. The findings highlight the need for gender-sensitive policies that promote earlier diagnosis and equitable treatment for women.

### 6.3 Cardiovascular Risk Factors in the Yemeni Population

Obesity remains relatively low in Yemen, with a prevalence of 8.8%, compared to much higher rates in Saudi Arabia (39.6%) and Egypt (46% in females). Cultural practices, food availability, and levels of physical activity likely explain this difference. Western countries, where obesity rates between men and women are more equal, benefit from shared societal roles and improved health awareness. In this study, diabetes was found in 31% of patients, which is lower than in Gulf countries (57–63%) and Egypt (43%), but higher than in Europe and Latin America (11–25%). Yemen’s lower rate may reflect a continued reliance on traditional diets and limited consumption of processed foods. Hypertension was present in 41% of cases, which is lower than in the Gulf (e.g., 65% in Saudi Arabia and 60.9% in the UAE) but close to rates in North Africa (45%). In contrast, Europe has much lower rates (15.7–19.2%), possibly due to more effective prevention and early intervention.

Smoking was reported in 61% of patients, much higher than in the Gulf (34%), Egypt (48.1%) (**13)**, and North Africa (14%) **(16, 17)**. Among these, 52.9% were both smokers and khat users, highlighting the compounding influence of cultural behaviors on cardiovascular risk. In contrast, only 14.8% of patients were neither smokers nor khat chewers. Khat chewing increases the desire to smoke more cigarettes and is commonly practiced in enclosed spaces, which may lead to elevated exposure to passive smoke. This combination further increases cardiovascular risk, especially among family members present during khat sessions.

These findings underscore the urgent need for culturally tailored prevention strategies, stronger emergency response systems, and equitable access to early cardiovascular diagnosis and intervention in Yemen.

### 6.4 Refined Discussion – Clinical Characteristics and Risk Profiles (Table 5.6)

Mean fasting blood glucose was 140 ± 71 mg/dL, random glucose 158 ± 92 mg/dL, systolic pressure 113 ± 25 mmHg, and diastolic 69.1 ± 15 mmHg—values lower than those reported in Saudi Arabia and the UAE **(18, 19)** potentially due to Yemen’s limited access to care and dependence on home-cooked food. Heart rate was higher (89 ± 20 bpm) than in the Gulf (84.6 bpm) and Europe (France 82.5 bpm, Portugal 59.7–65 bpm) **(5, 20, 21)** possibly due to widespread khat chewing and sympathetic stimulation. Lipid profiles were generally lower: total cholesterol 3.78 mmol/L, LDL 2.28 mmol/L, HDL 0.9 mmol/L, and triglycerides 1.8 mmol/L—well below levels in Saudi Arabia and Egypt **(22, 23)**, Still, only 4.9% had diagnosed dyslipidemia, compared to 82.7% in Saudi Arabia and 57% in Gulf countries (**16, 24)**, suggesting underdiagnosis. Typical chest pain was present in 69.7% of cases, with dyspnea frequent in patients ≥70 years. STEMI was the most common presentation (52.3%), surpassing Canada (40.7%) and Europe (42.5%) **(24, 25)**likely reflecting delayed care and a younger affected population.

ACS subtypes varied with age: STEMI declined from **40.4%** (≤50 years) to **16.7%** (>70 years), while NSTEMI and unstable angina increased with age (p < 0.005). This pattern aligns with global data showing atypical presentation in the elderly.

Past cardiac history was less common than in neighboring countries: CAD (24.5%), MI (22.2%), PCI (10.6%), CABG (2.7%)—versus higher rates in Saudi Arabia and Portugal **(21, 26)**.

In conclusion, the Yemeni ACS profile reflects a mix of region-specific risks (khat, smoking), socioeconomic constraints, and delays in care. These insights stress the need for preventive strategies and improved cardiac healthcare systems in Yemen.

### 6.5 Medical Therapy and Outcomes (Table 5.7)

Therapeutic interventions differed significantly by age. Patients >70 received more aspirin (93%), beta-blockers, ACE inhibitors, antibiotics, statins, and diuretics (p < 0.05). Clopidogrel and morphine were more common in the 51–70 age group, while metoprolol and digoxin were used more in younger patients. Only 18.1% of STEMI patients received thrombolysis, and 12.6% qualified for PPCI, reflecting significant gaps in reperfusion therapy.

Complications occurred in 42.8% of cases (n=191), with infections being most frequent (25%, p = 0.008), including sepsis and chest infections (7%, p = 0.000). Bleeding, renal failure, and conduction block affected 2–3% of patients. In-hospital mortality was 14.3%, while five-year mortality was 19.3%, considerably higher than in Saudi Arabia (7.9%) and Portugal (3.9%) **(21, 26)**. These outcomes likely stem from late presentation and inadequate infection control.

In contrast to the Netherlands, where 60% of cardiac patients access rehabilitation (reducing recurrence by 31% **(27)**, Yemen lacks structured programs and public health insurance, especially in government hospitals. This highlights the urgent need for improved emergency care, continuity of care, and a digital risk stratification system in facilities like the Military Cardiac Center in Sana’a.

### 7.1 Conclusion

This study revealed a critical gap in the management of STEMI patients in Yemen, with only 30.1% receiving any form of reperfusion therapy—12.6% via PCI and 18.1% via streptokinase. The majority (69.9%) did not receive reperfusion, likely contributing to the high in-hospital mortality rate of 14.3%, which surpasses regional benchmarks. These findings underscore severe systemic limitations, including inadequate access to interventional cardiology, delayed patient presentation, lack of emergency medical services, and the absence of universal health coverage. Addressing these gaps is essential to improve outcomes for acute cardiac patients in Yemen.

### 7.2 Recommendations

Based on the study findings, the following actions are recommended to address the burden of ACS in Yemen:

1. Launch public awareness campaigns targeting modifiable cardiovascular risk factors.
2. Introduce universal health insurance to improve equitable access to care.
3. Develop emergency medical transport systems for timely hospital access.
4. Offer free annual cardiac screenings for individuals aged >30, particularly in rural areas.
5. Guarantee free treatment for ACS in public hospitals.
6. Expand cardiac care services by establishing specialized centers nationwide.
7. Implement electronic health records and risk stratification tools for ongoing patient monitoring.

## Data Availability

The data supporting the findings of this study are available from the corresponding author upon reasonable request. The dataset will be made publicly available after the manuscript is published in a peer-reviewed journal.

## Appendix A: Declaration of English reviewer approval

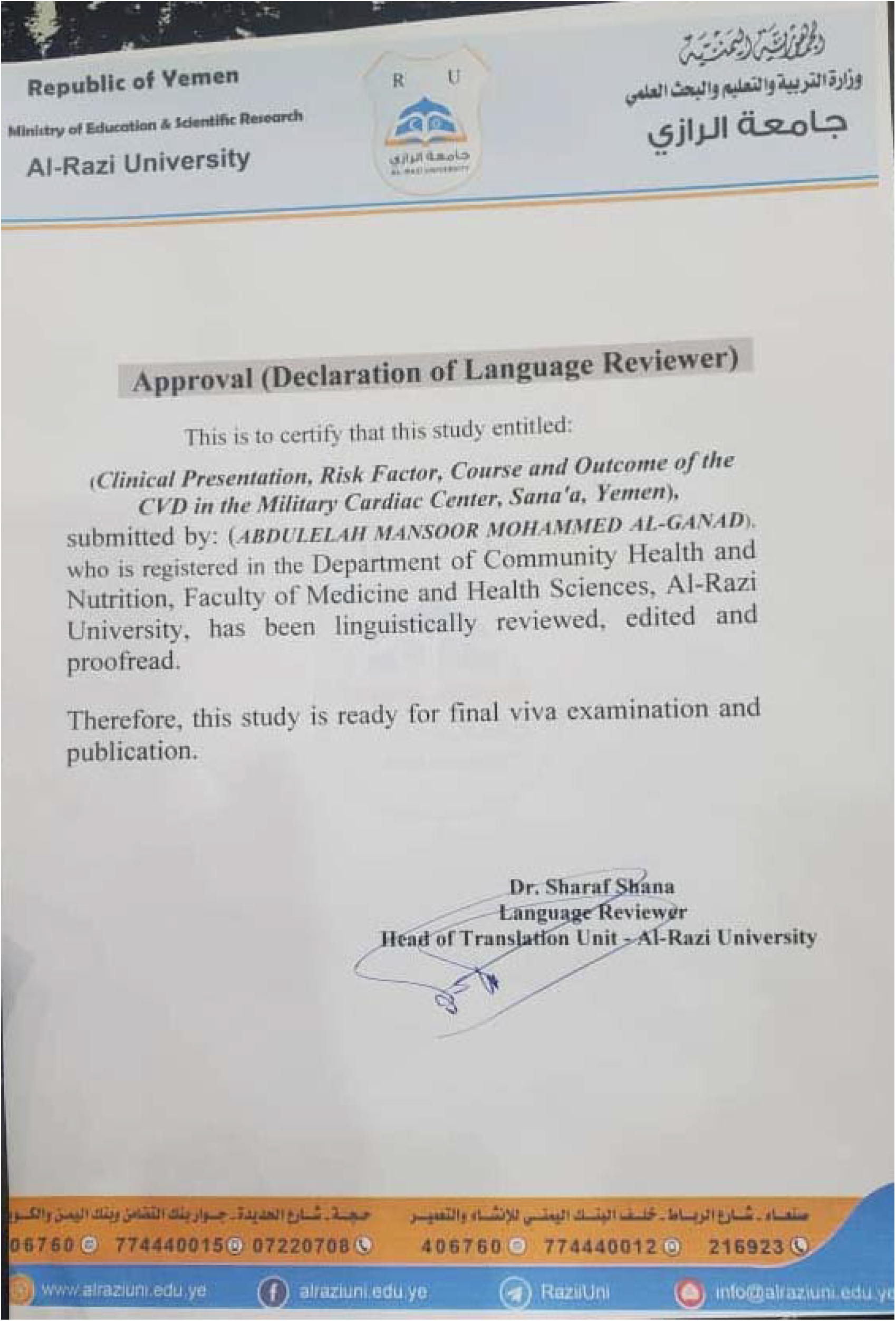

## Appendix D: Scientific Review Confirmation

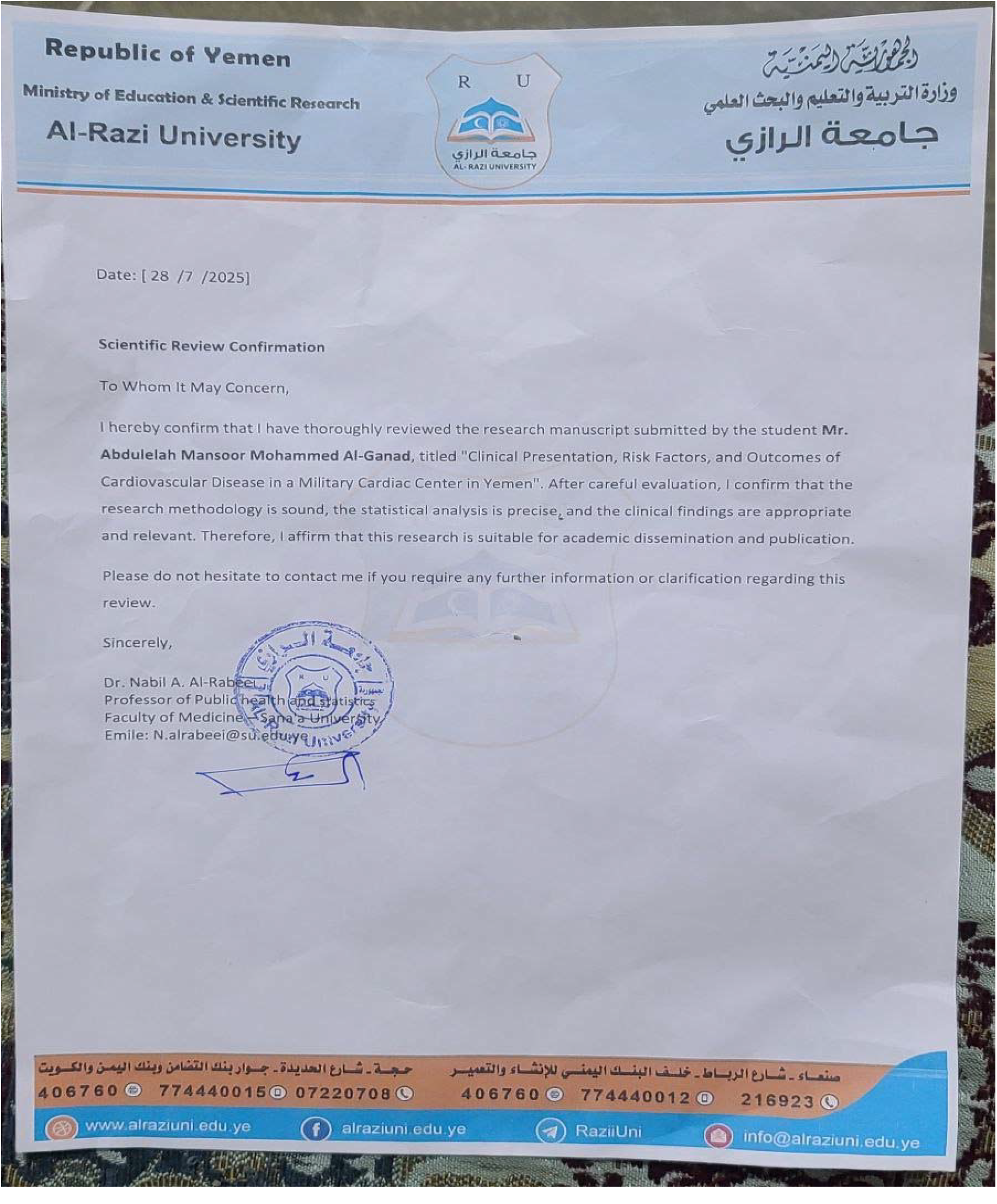

Appendices (Ethical Approvals and Institutional Permissions)

- Appendix B – Title Consent from Digital Library of the National Information Center This document confirms that the research was registered in the national digital repository. The original consent letter (in Arabic) is available upon request from the corresponding author.
- Appendix C – Military Cardiac Center Approval Formal institutional permission was obtained from the Military Cardiac Center in Sana’a, Yemen. The Arabic-language approval letter is available upon request from the corresponding author.
- Appendix E – Supervisory Letter to Prof. Ahmed Al-Mutarb This letter outlines the academic supervision and endorsement of the study. The original letter is written in Arabic and can be provided upon request. Note: All supporting Arabic documents have been removed from this version of the manuscript to comply with medRxiv’s policy. Please contact the corresponding author for access to these materials.

## Author Statement

This thesis was submitted by Abdulelah Mansoor Mohammed Al-Ganad in full fulfillment of the requirements for the Master’s degree in Epidemiology and Medical Biostatistics at Al-Razi University, Sana’a, Yemen.

## Acknowledgements

The authors gratefully acknowledge Dr. Ahmed Al-Motarreb, Dr. Taha Al-Maimoony, and Prof. Nabil Al-Rabeei for their invaluable supervision and review of this research. We also extend our thanks to Al-Razi University and the Military Cardiac Center for their institutional support and to the National Information Center for assisting with data access and record verification.

## Author Contributions

Abdulelah Mansoor Al-Ganad: Conceptualized and designed the study, developed the research methodology, collected clinical data, performed statistical analysis, interpreted the results, drafted the initial manuscript, formatted tables and figures, prepared the final version of the paper, and submitted the preprint. He is the corresponding author.

Prof. Ahmed Al-Motarreb: Professor of Cardiology and Head of the Cardiology Department, Faculty of Medicine – Sana’a University. He served as the principal academic supervisor for the entire study, providing continuous guidance throughout all phases of the research. His extensive expertise in cardiology was instrumental in shaping the study design, interpreting clinical data, and ensuring the scientific rigor of the study. He also offered critical feedback and a thorough manuscript review, significantly enhancing the quality and clarity of the final work.

Dr. Taha Al-Maimoony: Associate Professor of Cardiology – Sana’a University, and the external examiner of the master’s thesis. He provided valuable scientific comments that were integrated into the final published version.

Prof. Nabil Al-Rabeei: Professor of Public Health and Medical Biostatistics – Al-Razi University; internal supervisor of the master’s thesis. He supervised and reviewed the statistical analysis and contributed to improving the methodological quality.

Dr. Abdulaziz Mohammed Ali Aonallah: Assisted in collecting clinical data from patient records at the Military Cardiac Center in Sana’a.

Dr. Esmail Mohamed Sad Al-Dabis: Made a significant contribution to data analysis and quality assurance.

